# Identification of COVID-19 Subtypes Based on Immunogenomic Profiling

**DOI:** 10.1101/2021.01.24.21250387

**Authors:** Qiushi Feng, Xiaosheng Wang

**Affiliations:** Biomedical Informatics Research Lab, School of Basic Medicine and Clinical Pharmacy, China Pharmaceutical University, Nanjing 211198, China; Big Data Research Institute, China Pharmaceutical University, Nanjing 211198, China

**Keywords:** COVID-19 subtypes, antiviral immune response, gene expression profiles, clinical outcomes, clustering analysis

## Abstract

Although previous studies have shown that the host immune response is crucial in determining clinical outcomes in COVID-19 patients, the association between host immune signatures and COVID-19 patient outcomes remains unclear. Based on the enrichment levels of 11 immune signatures (eight immune-inciting and three immune-inhibiting signatures) in leukocytes of 100 COVID-19 patients, we identified three COVID-19 subtypes: Im-C1, Im-C2, and Im-C3, by clustering analysis. Im-C1 had the lowest immune-inciting signatures and high immune-inhibiting signatures. Im-C2 had medium immune-inciting signatures and high immune-inhibiting signatures. Im-C3 had the highest immune-inciting signatures while the lowest immune-inhibiting signatures. Im-C3 and Im-C1 displayed the best and worst clinical outcomes, respectively, suggesting that antiviral immune responses alleviated the severity of COVID-19 patients. We further demonstrated that the adaptive immune response had a stronger impact on COVID-19 outcomes than the innate immune response. The patients in Im-C3 were younger than those in Im-C1, indicating that younger persons have stronger antiviral immune responses than older persons. Nevertheless, we did not observe a significant association between sex and immune responses in COVID-19 patients. In addition, we found that the type II IFN response signature was an adverse prognostic factor for COVID-19. Our identification of COVID-19 immune subtypes has potential clinical implications for the management of COVID-19 patients.

## Background

The COVID-19 pandemic caused by SARS-CoV-2 has resulted in more than 36 million cases and one million deaths as of October 8, 2020 (*1*). The host immune defense system and immune response are crucial factors responsible for clinical outcomes in COVID-19 patients (*2-7*). For example, Takahashi et al. demonstrated that sex biases in COVID-19 were associated with differences in immune responses between males and females (*2*). The excessive immune response to SARS-CoV-2, known as COVID-19 cytokine storm, may cause severe COVID-19 (*8*). Despite these previous studies, the association between host immune responses and clinical outcomes in COVID-19 patients remains unclear.

In this study, using a publicly available RNA-Seq gene expression profiles in 100 leukocyte samples from COVID-19 patients (*9*), we performed an unsupervised learning to identify COVID-19 subtypes based on the enrichment levels of 11 immune signatures. Furthermore, we characterized the immunological and clinical features of these COVID-19 subtypes.

## Results

### Identification of COVID-19 subtypes based on immune signature enrichment levels

Based on the enrichment levels of 11 immune signatures, using the hierarchical clustering method, we identified three COVID-19 immune subtypes, termed Im-C1, Im-C2, and Im-C3 (Fig. 1A). Im-C1 had the lowest enrichment levels of HLA Class II, CD8+ T cells, Tfh, Th1 cells, Th2 cells, NK cells, pDCs, and cytolytic activity (termed immune-inciting signatures) while high enrichment levels of type II IFN response, Treg, and neutrophils (termed immune-inhibiting signatures). Im-C2 had medium immune-inciting signatures and high immune-inhibiting signatures. In contrast to Im-C1, Im-C3 had the highest immune-inciting signatures and the lowest immune-inhibiting signatures. Thus, Im-C3 and Im-C1 displayed the strongest and weakest antiviral immune responses, respectively. As expected, the ratios of immune-stimulatory to immune-inhibitory signatures (CD8+/CD4+ regulatory T cells and pro-/anti-inflammatory cytokines) were the highest in Im-C3 and the lowest in Im-C1 (one-tailed Mann–Whitney U test, *P* < 0.01) (Fig. 1B). Principal component analysis confirmed that these COVID-19 cases could be divided into three subgroups based on the ssGSEA scores of these immune signatures (Fig. 1C).

**Fig 1.**
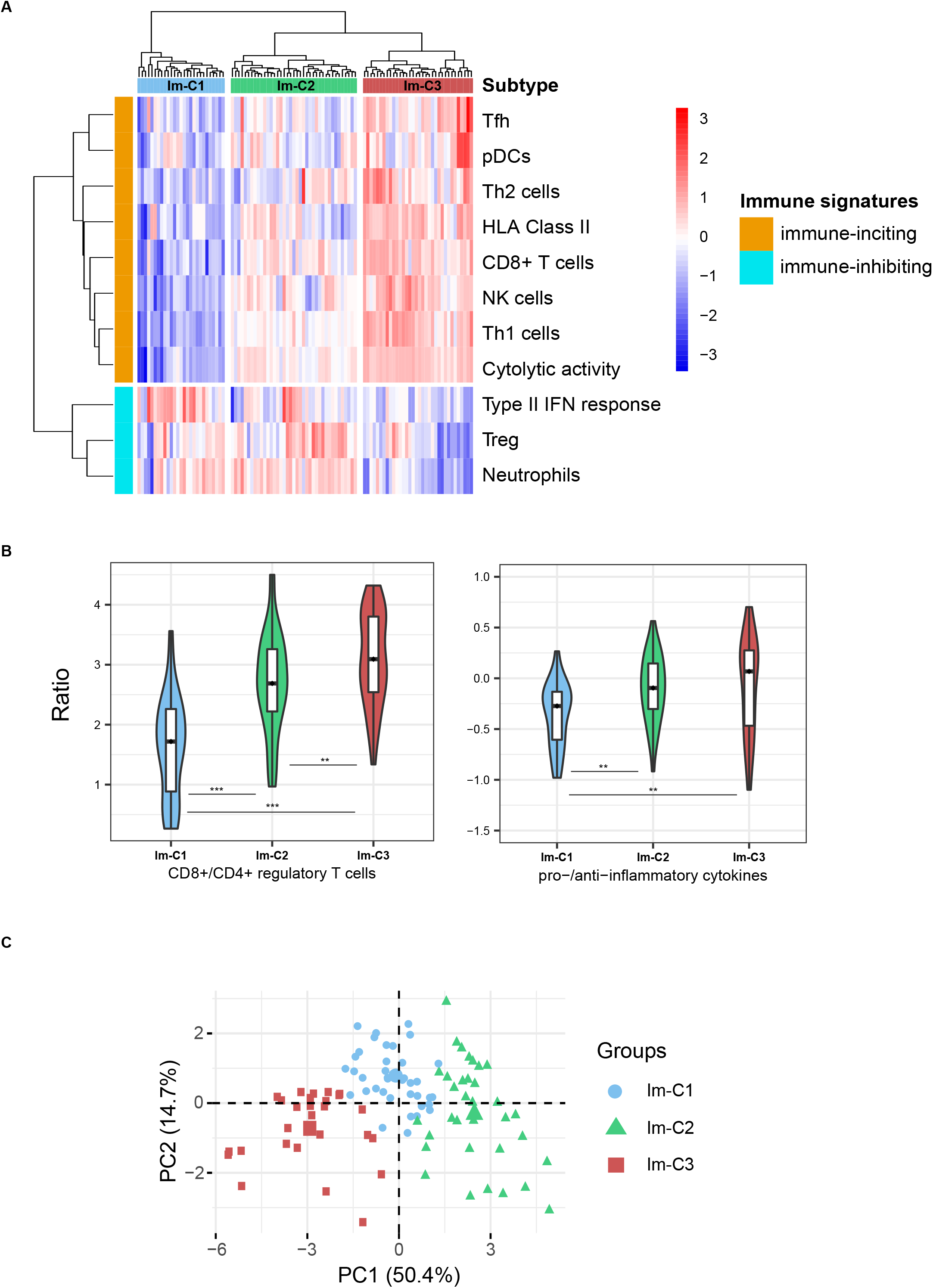
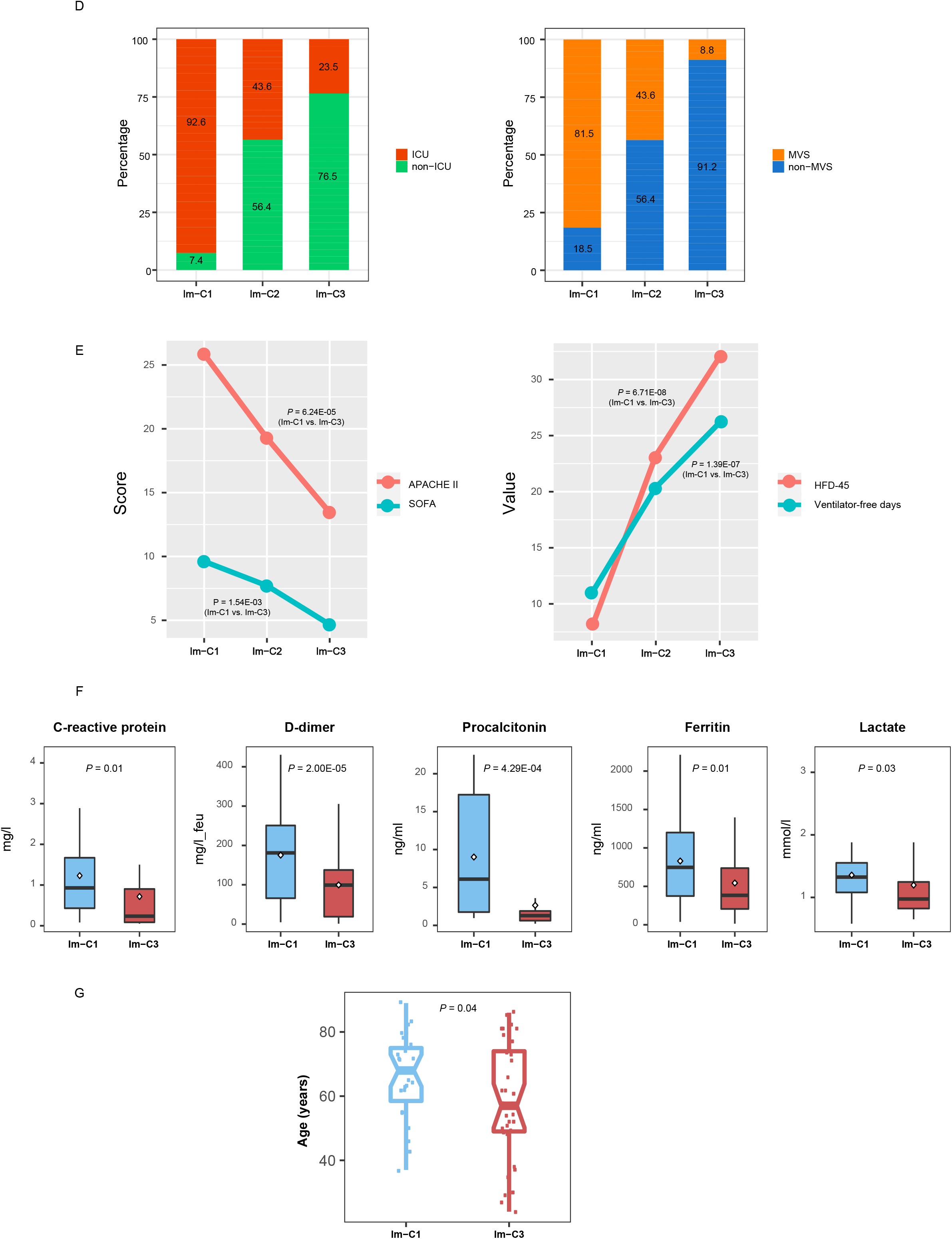
Identification of COVID-19 immune subtypes. **A**. Hierarchical clustering of 100 COVID-19 patients based on the enrichment scores of 11 immune signatures in leukocytes. **B**. Comparisons of the ratios of immune-stimulatory to immune-inhibitory signatures between the three COVID-19 subtypes. The ratios were the mean expression levels of the marker genes of immune-stimulatory signatures over those of immune-inhibitory signatures (log2-transformed). The Student’s *t* test *P*-values are indicated. * *P* < 0.05, ** *P* < 0.01, *** *P* < 0.001 (This also applies to the following figures). **C**. Principal component analysis confirming that COVID-19 can be divided into three subgroups based on the enrichment scores of the 11 immune signatures. **D**. Proportions of COVID-19 patients admitted to intensive care unit (ICU) or requiring mechanical ventilatory support (MVS) in the COVID-19 subtypes. **E**. Comparisons of the scores of the Acute Physiology and Chronic Health Evaluation (APACHE II) and the Sequential Organ Failure Assessment (SOFA), ventilator-free days, and the hospital-free days at day 45 (HFD-45) values between the three COVID-19 subtypes. **F**. Comparisons of the COVID-19 severity-associated laboratory measurements between COVID-19 subtypes. **G**. Comparisons of the age between COVID-19 subtypes. The one-tailed Mann–Whitney U test P-values are shown.

We found that the proportion of the COVID-19 patients admitted to intensive care unit (ICU) was the lowest in Im-C3 (23.5%) and the highest in Im-C1 (92.6%) (Fisher’s exact test, *P* < 0.001) (Fig. 1D). Moreover, the proportion of COVID-19 patients requiring mechanical ventilatory support (MVS) was the lowest in Im-C3 (8.8%) and the highest in Im-C1 (81.5%) (Fisher’s exact test, *P* < 0.001). The scores of the Acute Physiology and Chronic Health Evaluation (APACHE II) and the Sequential Organ Failure Assessment (SOFA), both of which measure the severity of ICU patients (*10*), were significantly different between the three subtypes: Im-C3 < Im-C2 < Im-C1 (one-tailed Mann–Whitney U test, *P* < 0.001) (Fig. 1E). The numbers of ventilator-free days, which is an outcome measure in treatments for acute respiratory distress syndrome (*11*), and the hospital-free days at day 45 (HFD-45) values, which correlated inversely with disease severity, were significantly different between the three subtypes: Im-C3 > Im-C2 > Im-C1 (one-tailed Mann–Whitney U test, *P* < 0.001) (Fig. 1E). Additionally, some laboratory measurements, such as C-reactive protein, D-dimer, procalcitonin, ferritin, and lactate, whose elevation was associated with COVID-19 severity, tended to display the lowest levels in Im-C3 while the highest levels in Im-C1 (Fig. 1F). Altogether, these results showed that the Im-C3 subtype of COVID-19 had the best outcomes, while the Im-C1 subtype had the worst outcomes. It suggests that antiviral immune responses can reduce COVID-19 disease severity. Furthermore, we found that the patients in Im-C3 were younger than those in Im-C1 (one-tailed Mann–Whitney U test, *P* = 0.04) (Fig. 1G). It indicates that younger persons tended to have a stronger antiviral immune response than older persons after SARS-CoV-2 infection. However, we did not observe a significant difference in the proportions of female and male patients between Im-C3 and Im-C1 (Fisher’s exact test, *P* = 0.61).

### Associations between immune signatures and clinical features in COVID-19 patients

We further analyzed associations between immune signatures and clinical features in COVID-19 patients. As expected, the elevated enrichment of the eight immune-inciting signatures were correlated with higher HFD-45 values and more ventilator-free days (Spearman’s correlation test, *P* < 0.01, ρ > 0.3) (Fig. 2A). Moreover, their enrichment levels were significantly higher in non-ICU versus ICU patients and in non-MVS versus MVS patients (one-tailed Mann–Whitney U test, *P* < 0.01) (Fig. 2B). Collectively, these results indicate that the enrichment of these immune-inciting signatures has a positive association with outcomes in COVID-19 patients. In addition, we found five immune-inciting signatures (HLA Class II, CD8+ T cells, Th1 cells, Th2 cells, and NK cells) whose enrichment levels correlated inversely with ages of COVID-19 patients (*P* < 0.1, ρ < −0.18) (Fig. 2C). However, none of the eight immune-inciting signatures showed significantly different enrichment levels between female and male patients. Again, these results indicate that younger patients have a stronger immune response to SARS-CoV-2 infection than older patients, while the strength of immune response is not different between female and male patients. We further demonstrated the significant negative correlation between the immune-inciting signatures and ages of COVID-19 patients in two other RNA-Seq gene expression profiling datasets for COVID-19 patients (GSE156063 (*12*) and GSE152075 (*13*) (Fig. 2C). Likewise, the associations between gender and these immune-inciting signatures were not significant in both datasets. Interestingly, in GSE156063, six immune-inciting signatures displayed significantly lower enrichment levels in COVID-19 patients than in the patients infected with other viruses (one-tailed Mann–Whitney U test, *P* < 0.01) (Fig. 2D). It suggests that SARS-CoV-2 causes a weaker human host immune response compared to other viruses, a potential explanation for the higher infectivity and pathogenicity of SARS-CoV-2 versus other viruses. In contrast to the immune-inciting signatures, the immune-inhibiting signatures were likely to have a negative correlation with outcomes in COVID-19 patients, as evidenced by that the three immune-inhibiting signatures (type II IFN response, Treg, and neutrophils) displayed significantly higher enrichment levels in MVS than in non-MVS patients (one-tailed Mann–Whitney U test, *P* < 0.05) (Fig. 2E). In addition, the type II IFN response signature had inverse correlations with ventilator-free days and HFD-45 values (*P* < 0.05, ρ < −0.21) and was significantly higher in ICU versus non-ICU patients (Fig. 2F).

**Fig 2.**
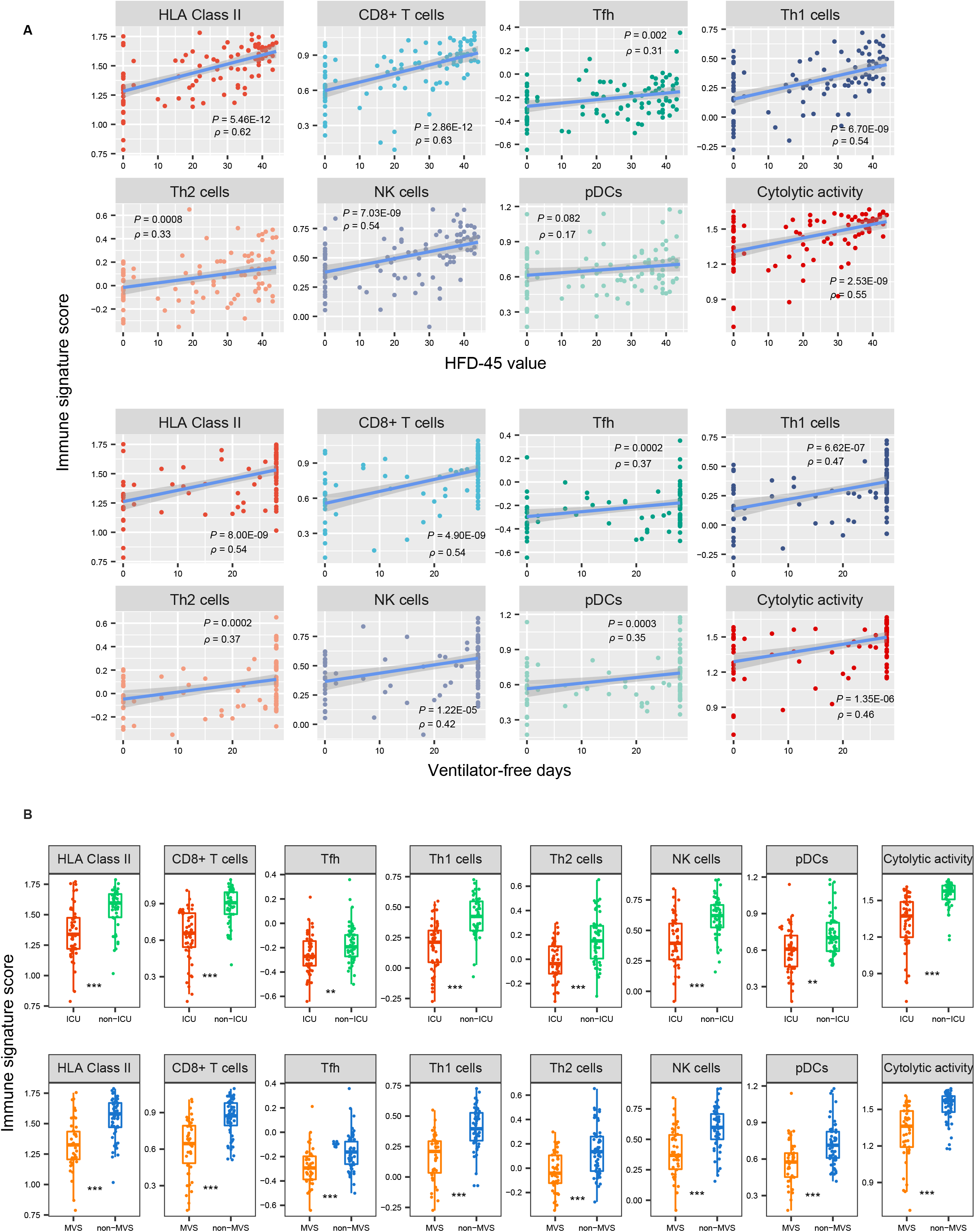
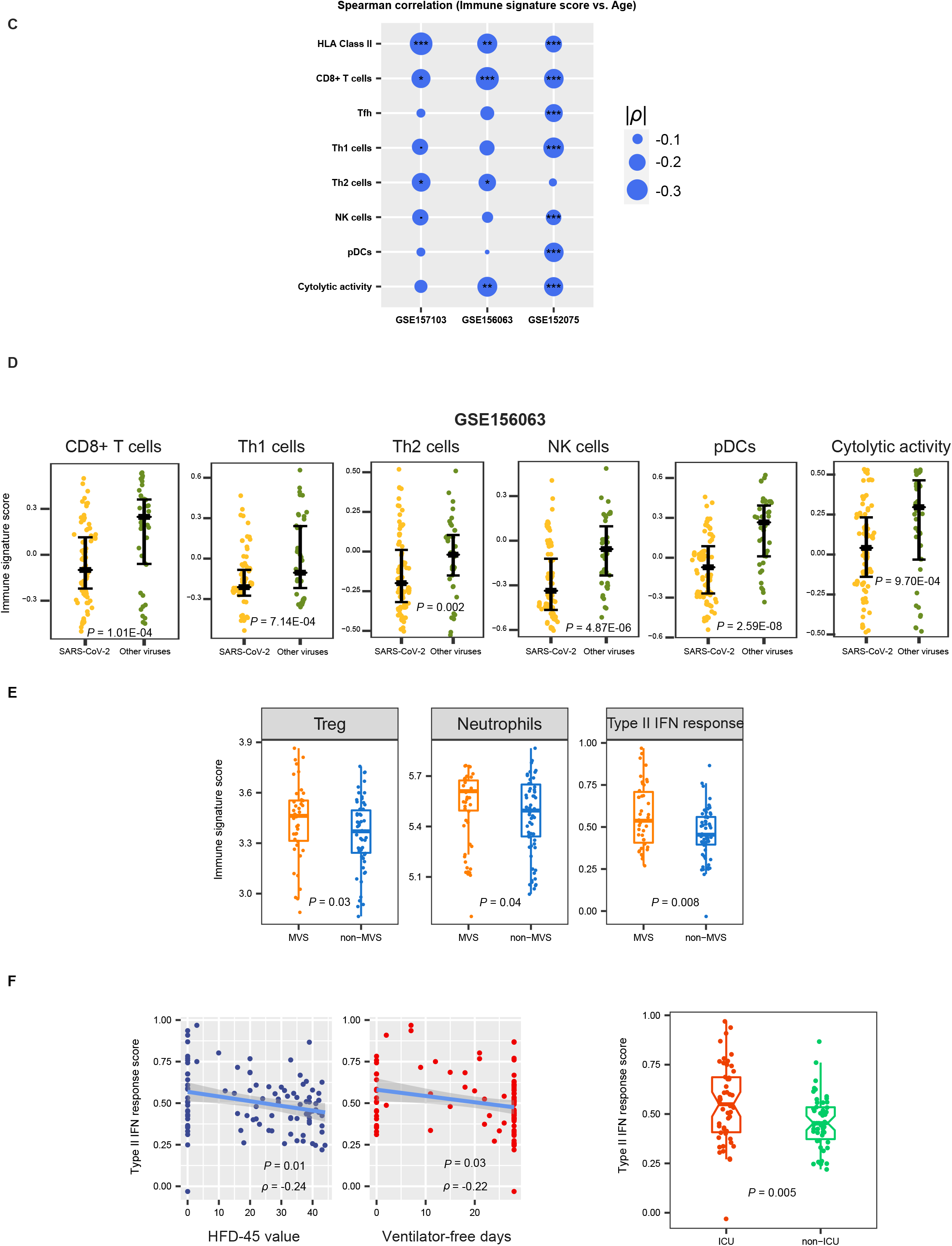
Associations between immune signatures and clinical features in COVID-19 patients. **A**. Associations between immune signature scores and HFD-45 values and ventilator-free days. The Spearman’s correlation test *P*-values and correlation coefficients are shown. **B**. Comparisons of immune signature scores between ICU and non-ICU patients and between MVS and non-MVS patients. The one-tailed Mann– Whitney U test *P*-values are shown. **C**. Associations between immune signature scores and age. **D**. Comparisons of immune signature scores between COVID-19 patients and the patients infected with other viruses. **E**. Comparisons of the scores of three immune-inhibiting signatures (type II IFN response, Treg, and neutrophils) between non-MVS and MVS patients. **F**. The scores of type II IFN response have negative correlations with HFD-45 values and ventilator-free days and are higher in ICU than in non-ICU patients.

### Comparison of the contribution of different factors in the prediction of COVID-19 outcomes

To compare the contribution of different factors in the prediction of outcomes in COVID-19 patients, we used the logistic regression model with five predictors (age, gender, CD8+ T cell score, NK cell score, and type II IFN response score) to predict ICU (= 1) versus non-ICU (= 0) and MVS (= 1) versus non-MVS (= 0) patients, respectively. In predicting MVS versus non-MVS, age, CD8+ T cells, and NK cells were significant negative predictors, while type II IFN response was a significant positive predictor (*P* < 0.05) (Fig. 3). In predicting ICU versus non-ICU, CD8+ T cells and NK cells were significant negative predictors (*P* < 0.1), while type II IFN response was a positive predictor (*P* = 0.186, β = 1.98). These results indicate that COVID-19 outcomes are correlated positively with immune-inciting signatures and negatively with immune-inhibiting signatures and age, consistent with previous results. Meanwhile, logistic regression analyses indicate that the adaptive immune response (CD8+ T cells) has a stronger impact on COVID-19 outcomes than the innate immune response (NK cells), as evidenced by the larger β values of CD8+ T cells versus NK cells in predicting ICU and MVS.

**Fig 3.**
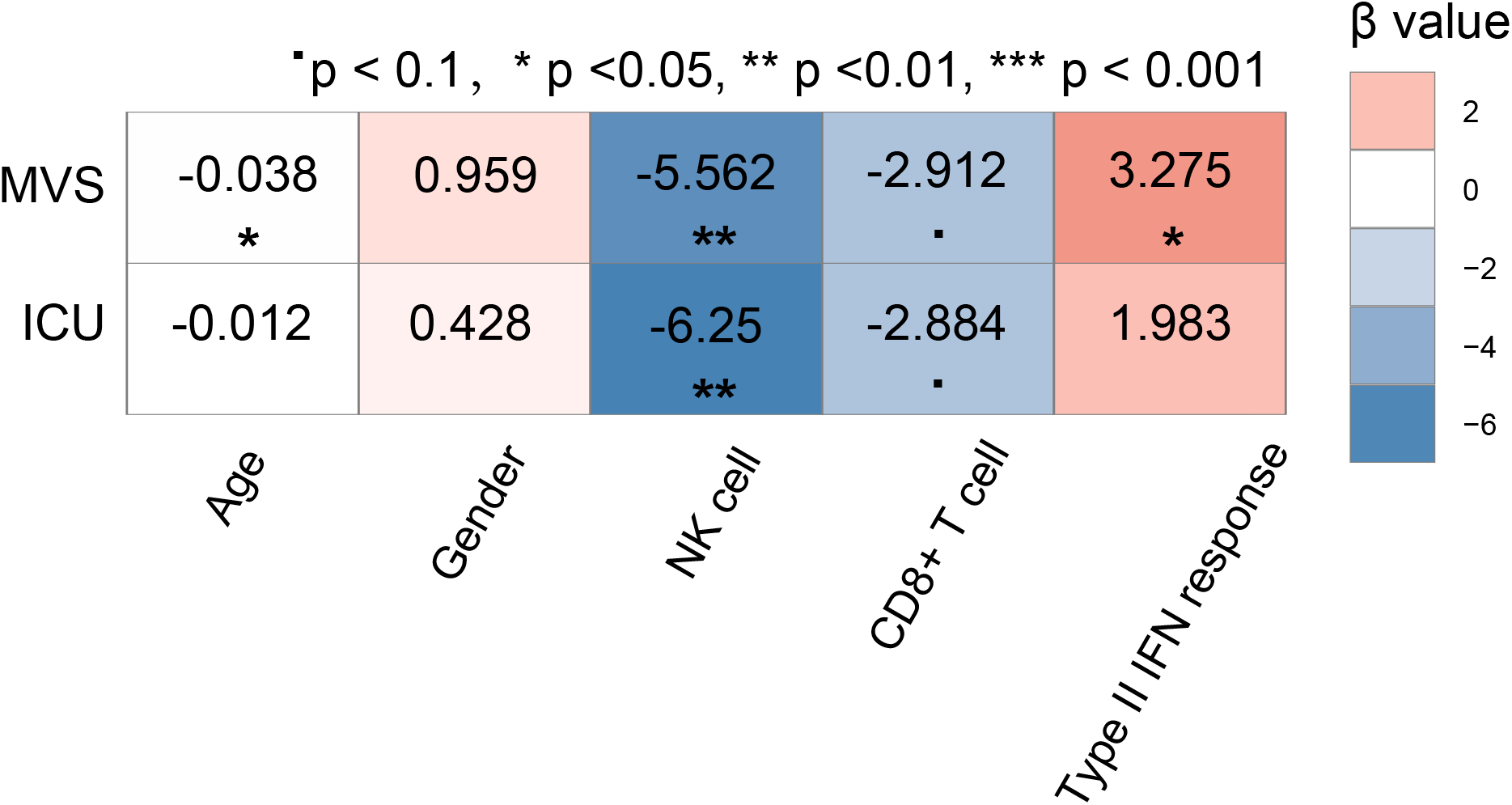
Prediction of ICU versus non-ICU and MVS versus non-MVS patients using five predictors (age, gender, CD8+ T cell score, NK cell score, and type II IFN response score) by logistic regression analyses. The standardized regression coefficients (β values) are shown.

### Identification of gene ontology differentially enriched between COVID-19 subtypes

WGCNA identified seven gene modules (indicated in cyan, light yellow, brown, light green, magenta, black, and turquoise color, respectively) that significantly differentiated COVID-19 patients by COVID-19 subtypes (Im-C1, Im-C2, and Im-C3) and outcomes (HFD-45, ICU, and MVS) (Fig. 4). The representative gene ontology (GO) terms associated with the gene modules highly enriched in Im-C3 while lowly enriched in Im-C1 included viral transcription, mitochondrial protein complex, RNA processing, and endoplasmic reticulum part. Consistently, these modules were associated with better outcomes of COVID-19. Besides, the immune response downregulated in Im-C1 was positively associated with COVID-19 outcomes. In addition, the protein modification process representing the black module, which was downregulated in Im-C3 while upregulated in Im-C2, correlated with worse outcome of MVS.

**Fig 4.**
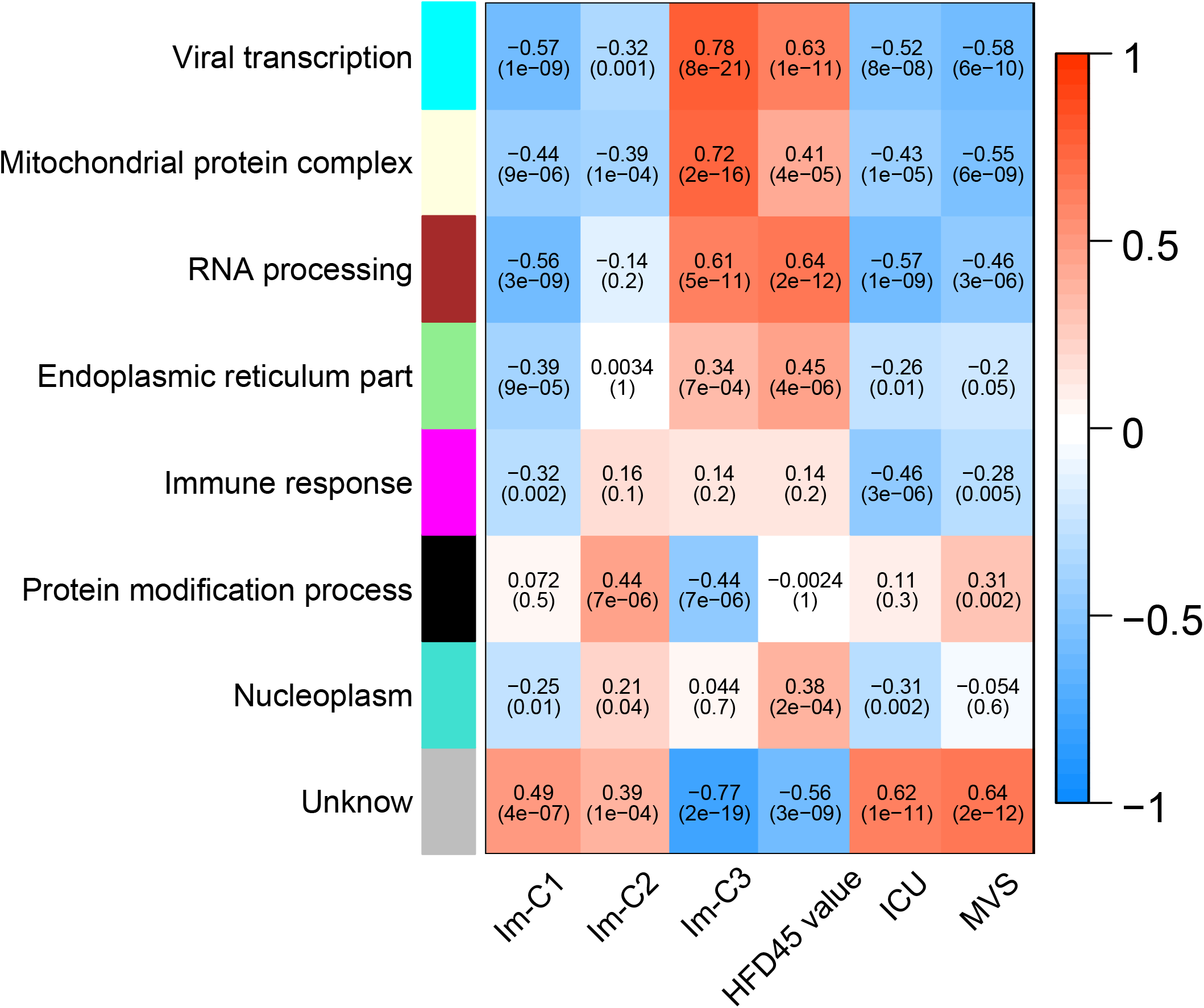
WGCNA identified seven gene modules significantly differentiating COVID-19 patients by COVID-19 subtypes and outcomes. The representative gene ontology terms for gene modules, correlation coefficients, and *P*-values are shown.

### Identification of genes and pathways differentially expressed between ICU and non-ICU COVID-19 patients

We identified 67 and 309 genes upregulated and downregulated in ICU versus non-ICU COVID-19 patients (Fig. 5A and Supplementary Table S1). We found a number of immune-related pathways associated with the upregulated genes in non-ICU, including antigen processing and presentation, natural killer cell mediated cytotoxicity, hematopoietic cell lineage, intestinal immune network for IgA production, T cell receptor signaling, cytokine-cytokine receptor interaction, chemokine signaling, Toll-like receptor signaling, RIG-I-like receptor signaling, cytosolic DNA-sensing, Jak-STAT signaling, NOD-like receptor signaling, and Fc epsilon RI signaling (Fig. 5B). Again, these results indicate the stronger immune response in non-ICU versus ICU COVID-19 patients. Furthermore, we performed a prediction of ICU versus non-ICU patients based on gene expression profiles in leukocyte samples from 100 COVID-19 patients (GSE157103). The 3-fold cross validation (CV) accuracy was 83.1%, and the area under the ROC curve (AUC) was 91.5% (Fig. 5C). It indicates that the gene expression profiles in leukocytes of COVID-19 patients could be a potentially useful predictor for the severity of COVID-19.

**Fig 5.**
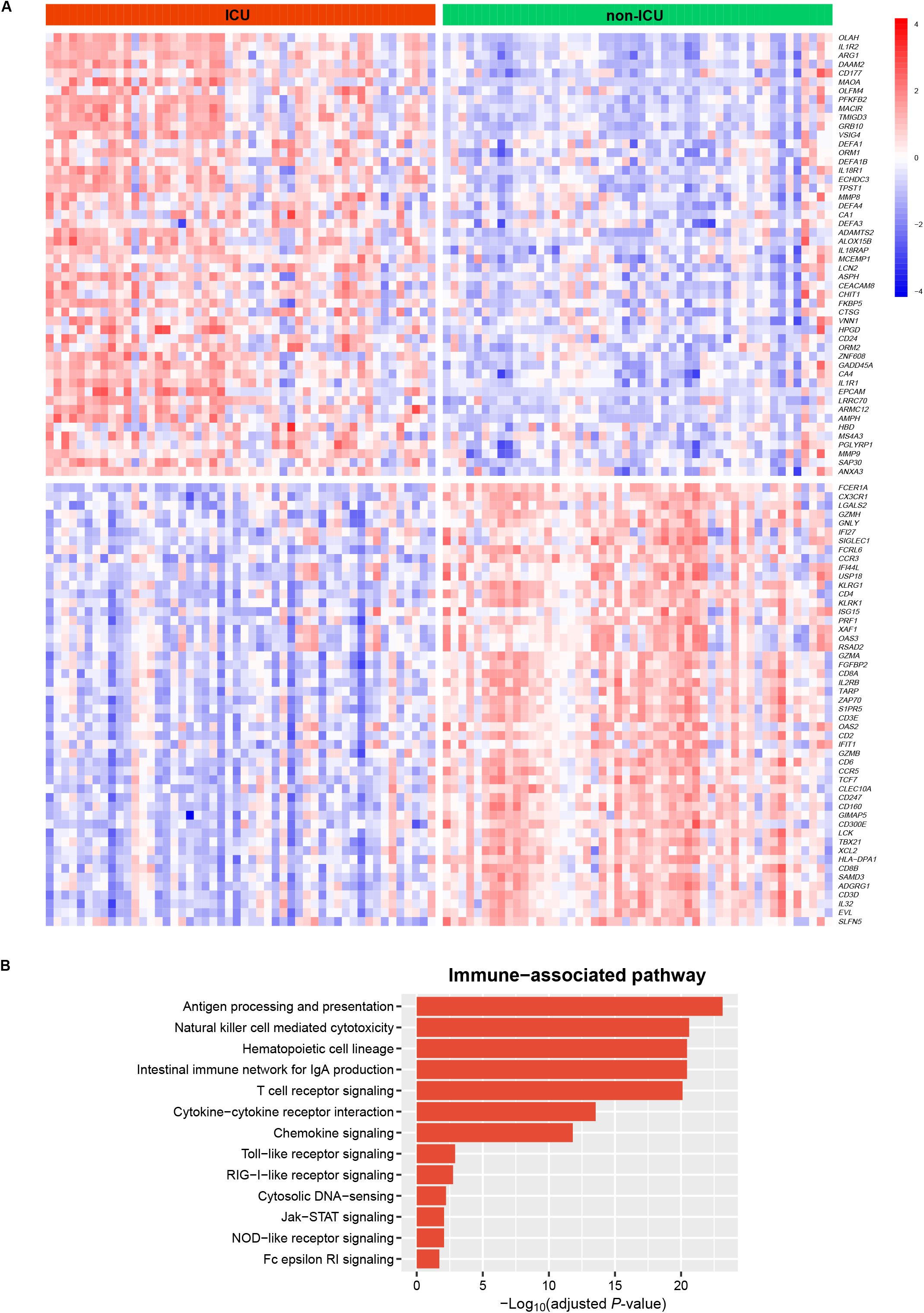
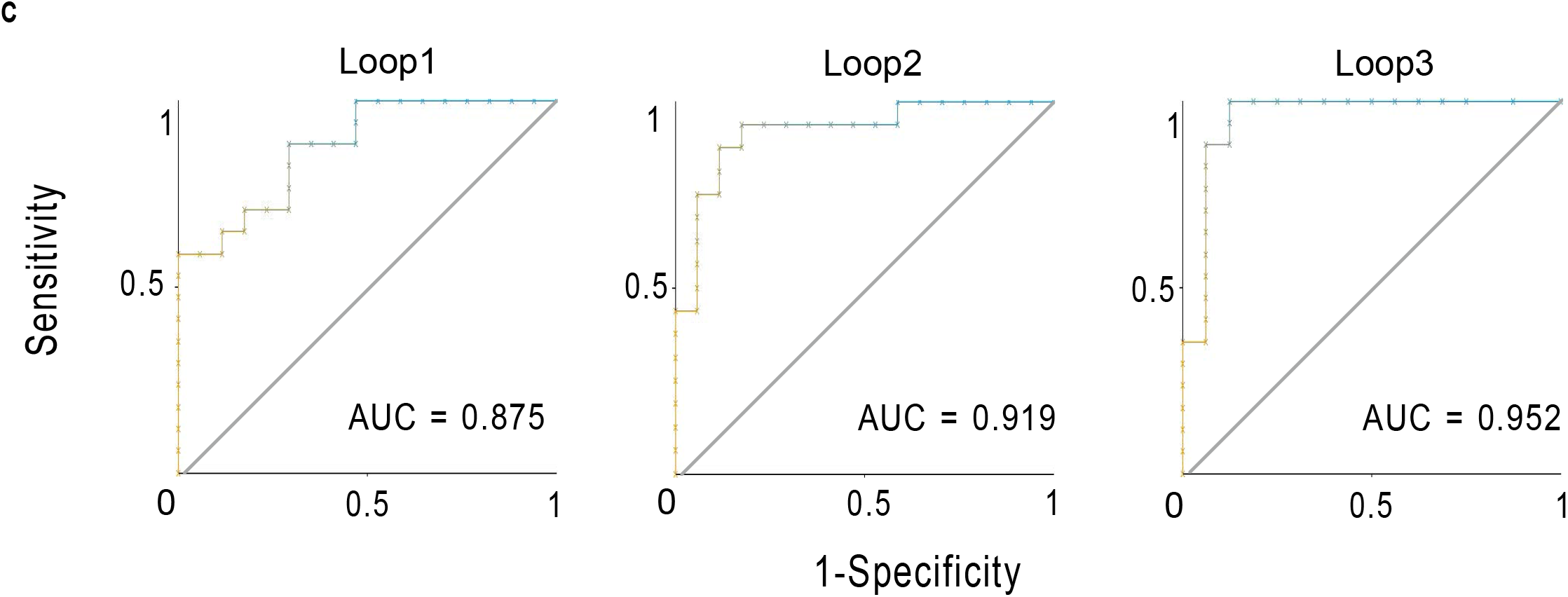
Genes and pathways differentially expressed between ICU and non-ICU COVID-19 patients. **A**. Heatmap for 50 and 50 genes showing the highest increases and decreases of the fold change of mean expression levels in ICU versus non-ICU patients, respectively. **B**. Immune-related pathways upregulated in ICU versus non-ICU patients. **C**. Prediction performance of gene expression profiles in leukocyte samples from 100 COVID-19 patients in the prediction of ICU versus non-ICU patients by Random Forest. The area under the ROC curve (AUC) is shown for each loop of the 3-fold cross validation (CV).

**Fig 6.**
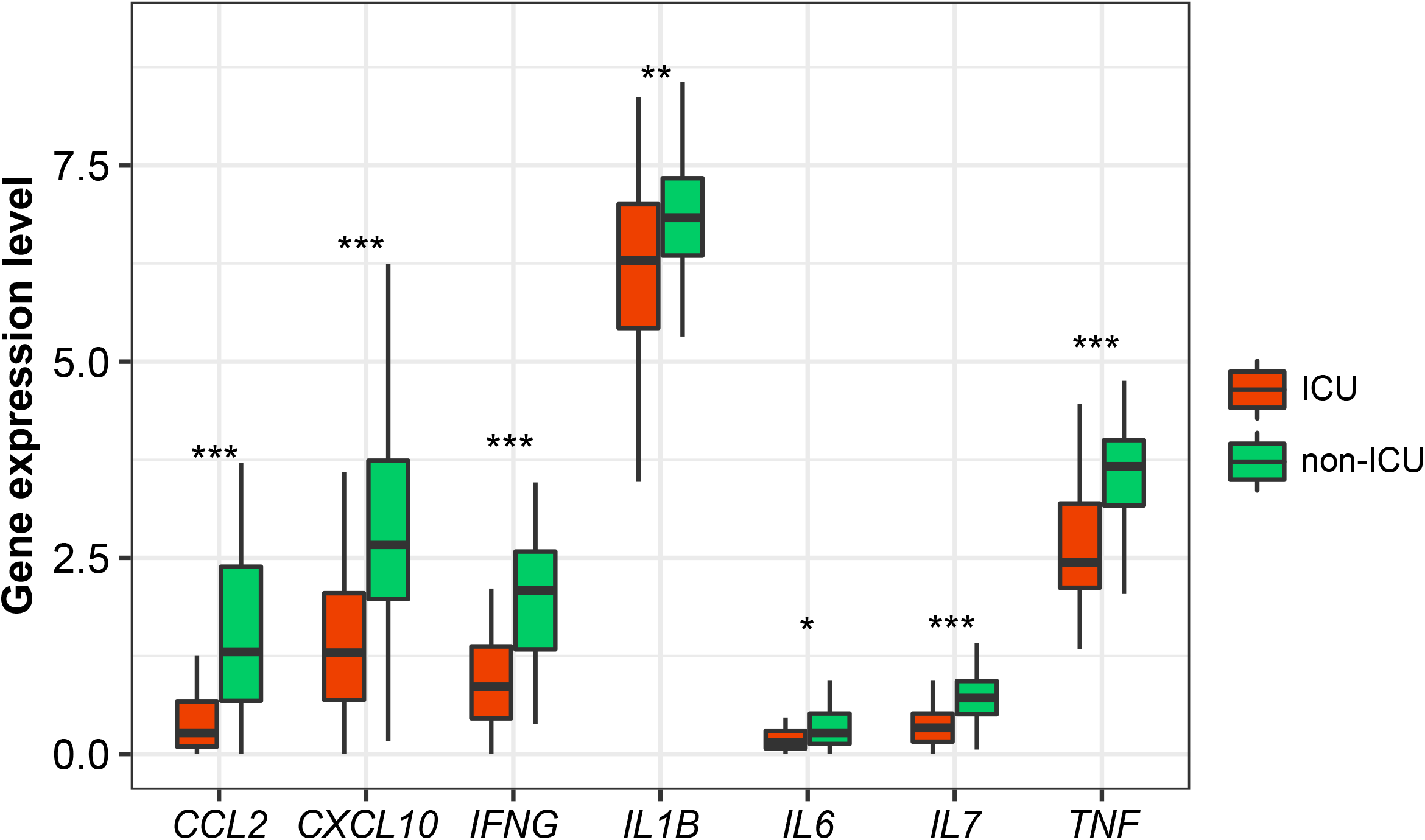
Comparisons of the expression levels of cytokine genes in leukocytes between ICU and non-ICU COVID-19 patients. The Student’s *t* test *P*-values are shown.

## Discussion

Based on the enrichment levels (ssGSEA scores) of 11 immune signatures in leukocytes of COVID-19 patients, we identified three COVID-19 subtypes: Im-C1, Im-C2, and Im-C3, by clustering analysis. The ssGSEA scores-based clustering method has been shown to be more robust than the gene expression values-based method in identifying subtypes of diseases (*14-16*). Im-C1 had the lowest immune-inciting signatures and high immune-inhibiting signatures. Im-C2 had medium immune-inciting signatures and high immune-inhibiting signatures. Im-C3 had the highest immune-inciting signatures while the lowest immune-inhibiting signatures. Im-C3 and Im-C1 COVID-19 patients had the best and worst clinical outcomes, respectively, suggesting that antiviral immune responses alleviated the severity of COVID-19 patients. We further demonstrated that the adaptive immune response exerted a greater impact on COVID-19 outcomes than the innate immune response. The patients in Im-C3 were younger than those in Im-C1, indicating that younger persons have stronger antiviral immune responses than older persons. Nevertheless, we did not observe a significant association between sex and immune responses in COVID-19 patients. In addition, we found that the type II IFN response signature was an adverse prognostic factor for COVID-19 in the dataset GSE157103. This result appears inconsistent with previous findings (1*7-18*). The reason behind this needs to be further investigated.

Our data suggest that a strong antiviral immune response can reduce COVID-19 severity. Thus, a strong host immune system is crucial for fighting against COVID-19, as bolstered by a recent study (1*9*). However, previous studies have revealed that serum inflammatory cytokine levels had an inverse association with clinical outcomes in COVID-19 patients (*20-23*). It suggests that excessive immune response, known as cytokine storm, may cause immunopathological damage in COVID-19 patients (*7*). We compared the expression levels of several cytokine genes in leukocytes between ICU and non-ICU COVID-19 patients, including *IL-6, IL-1*β, *TNF, CCL2, CXCL10, IFNG, IL7*. We found that these genes displayed significantly higher expression levels in non-ICU than in ICU patients (Fig. 5). A potential explanation for these different results could be the different sources of these cytokine.

## Methods

### Datasets

We downloaded the RNA-Seq gene expression profile datasets in leukocyte samples from 100 COVID-19 patients (GSE157103) and in SARS-CoV-2-infected human tissues from nasopharyngeal swabs (GSE152075 and GSE156063) from the Gene Expression Omnibus (GEO) (https://www.ncbi.nlm.nih.gov/geo/). Supplementary Table S2 summarizes these datasets.

### Quantification of the enrichment levels of immune signatures

We used the single-sample gene-set enrichment analysis (ssGSEA) score [24] to evaluate the enrichment level of an immune signature in a COVID-19 patient based on the gene expression profiles. The ssGSEA score represents the enrichment score of a gene set in a sample based on the degree of the genes in the gene set coordinately up- or down-regulated in the sample. We analyzed 11 immune signatures, including HLA Class II, CD8+ T cells, Tfh, Th1 cells. The gene sets representing these immune signatures are listed in Supplementary Table S3.

### Clustering analysis

We hierarchically clustered 100 leukocyte samples from COVID-19 patients (GSE157103) based on the ssGSEA scores of the 11 immune cell types. We performed the clustering analysis by using the R function “hclust” for hierarchical agglomerative clustering.

### Logistic regression analysis

We used logistic regression with five predictors (age, gender, CD8+ T cell score, NK cell score, and type II IFN response score) to predict ICU and MVS, respectively. The logistic regression analysis utilized the R function “glm” to fit the binary model and the R function “lm.beta” in the R package “QuantPsyc” to calculate the standardized regression coefficients (β values).

### Identification of gene ontology associated with COVID-19 subtypes

We used WGCNA (*25*) to identify the gene modules differentially enriched in COVID-19 subtypes and outcomes. The representative gene ontology (GO) terms associated with the gene modules were identified. The WGCNA analysis was performed by using the R package “WGCNA” (version 1.68).

### Pathway analysis

We identified differentially expressed genes between ICU and non-ICU COVID-19 patients using Student’s *t* test with a threshold of adjusted *P*-value (false discovery rate (FDR) < 0.05 and fold change (FC) of mean expression levels > 2. Based on the differentially expressed genes, we identified KEGG (*26*) pathways differentially enriched between ICU and non-ICU COVID-19 patients by (*27*) with a threshold of FDR < 0.05. The FDR was calculated by using the Benjamini-Hochberg method (*28*).

### Class prediction

We predicted ICU versus non-ICU patients based on gene expression profiles in leukocyte samples from 100 COVID-19 patients (GSE157103). We performed 3-fold CV in the 100 samples. Within each loop of the CV, we selected the 100 genes with the largest absolute t-scores in the comparison of ICU versus non-ICU patients in the training set; based on the 100 genes, we trained the Random Forest (RF) classifier and predicted ICU versus non-ICU patients in the test set. We reported the prediction performance (accuracy and AUC) as the average of them in the 3-fold CV. We carried out the prediction algorithm in Weka (*29*) with the number of trees in the RF set to 500.

### Statistical analysis

In comparison of two classes of data, we used Mann–Whitney U test if they were not normally distributed and used Student’s *t* test if they were normally distributed. We used Spearman’s correlation test to evaluate the correlation between two groups of data on the assumption that they were not normally distributed. We used Fisher’s exact test to evaluate the association between two categorical variables. All statistical analyses were performed in the R programming environment (version 4.0.2).

## Data Availability

In this study, using a publicly available RNA-Seq gene expression profiles in 100 leukocyte samples from COVID-19 patients GEO (https://www.ncbi.nlm.nih.gov/geo/)

## List of abbreviations

HLA: Human leukocyte antigen
Tfh: 
T follicular helper cells
Th1 cells: 
Type 1 T helper cells
Th2 cells: 
Type 2 T helper cells
NK: Natural killer
pDCs: 
Plasmacytoid dendritic cells
Treg: 
Regulatory T cells
ICU: 
Intensive care unit
MVS: 
Mechanical ventilatory support
HFD: 
Hospital-free days at day 45
WGCNA: 
Weighted gene co-expression network analysis
FDR: 
False discovery rate
RF: 
Random forest

## Declarations

### Ethics approval and consent to participate

Ethical approval was waived since we used only publicly available data and materials in this study.

### Consent for publication

Not applicable.

### Authors’ Contributions

QF performed data analyses and helped prepare for the manuscript. XW conceived of the research, designed the methods, and wrote the manuscript. All authors read and approved the final manuscript.

## Acknowledgments

Not applicable.

## Supplementary materials

Table S1. The genes differentially expressed between non-ICU and ICU COVID-19 patients.

Table S2. A summary of the datasets used in this study. Table S3. The gene sets representing immune signatures.

